# Circulating argonaute-bound microRNA-126 reports vascular dysfunction & response to treatment in acute & chronic kidney disease

**DOI:** 10.1101/2020.07.14.20153353

**Authors:** Kathleen M Scullion, A D Bastiaan Vliegenthart, Laura Rivoli, Wilna Oosthuyzen, Tariq E Farrah, Alicja Czopek, David J Webb, Robert W Hunter, Matthew A Bailey, Neeraj Dhaun, James W Dear

## Abstract

**Background:** Vascular and kidney dysfunction commonly co-exist. There is an unmet need for biomarkers of vascular health. Circulating microRNAs (miRs) are disease biomarkers; miR-126 is endothelial cell-enriched. We measured circulating miR-126 in rats with nephrotoxic nephritis (NTN) and humans with acute endothelial and renal injury (vasculitis associated with autoantibodies to neutrophil cytoplasm antigens (ANCA)). We then compared these findings to those from patients with chronic kidney disease (CKD) and end-stage renal disease (ESRD) and explored the relationship between miR-126 and markers of vascular dysfunction.

**Methods:** In rats, NTN was induced with nephrotoxic serum. Samples were prospectively collected from 70 patients with ANCA vasculitis (at presentation and once in treatment-induced remission), healthy subjects (n=60), patients with CKD (n=30) and ESRD before and after dialysis (n=15). In humans and rats, miR-126 and miR-122 (liver-specific control) were measured and correlated with vascular function.

**Results:** In NTN rats, miR-126 was selectively reduced. In ANCA vasculitis, pre-treatment circulating miR-126 was reduced compared to health (88-fold reduction, ROC-AUC 0.87 (0.80-0.94)). miR-126 increased 3.4-fold post-treatment but remained lower than in health (∼26-fold). There was no change in miR-122. Plasma fractionation demonstrated that argonaute 2-bound miR-126 increased with treatment of ANCA vasculitis. Urinary miR-126 decreased post-treatment. miR-126 did not differ between CKD and health but its concentration significantly correlated with measures of endothelial dysfunction. miR-126 was substantially reduced in ESRD (∼350 fold compared to health and CKD).

**Conclusions:** miR-126 may be a marker of vascular inflammation and has the potential to aid in clinical decision-making.

## Introduction

Vascular dysfunction commonly co-exists with kidney disease and contributes to an increased risk of cardiovascular disease (CVD). A severe, acute form of this vascular-renal phenotype is seen in patients presenting with anti-neutrophil cytoplasm antibody-associated vasculitis (AAV), a rare autoimmune disorder. The most frequent severe manifestations of AAV involve endothelial injury giving rise to a rapidly progressive glomerulonephritis and pulmonary hemorrhage. Despite current treatments overall survival remains poor^1, 2^ with many patients suffering chronic inflammation, a major contributor to the development and progression of both CVD^3^ and chronic kidney disease (CKD).^4^ Furthermore, those who respond to treatment remain at risk of further disease relapses.^5^

Identifying AAV early and assessing its response to treatment remain important clinical challenges. In those with renal involvement, the measurement of renal function using serum creatinine is often inadequate because substantial renal damage can occur before function is impaired to a detectable extent.^6^ Additionally, although serum creatinine may fall with treatment, it remains unclear whether histological inflammation continues once renal function has stabilized. Also, currently, there are no good measures of disease activity in those with extra-renal AAV alone. Biomarkers specific to small vessel inflammation would not only allow early implementation of appropriate treatments but also help identify those patients with grumbling disease activity and potentially predict disease relapses.

While AAV might be considered a phenotypic extreme, patients with CKD due to any cause are at an increased risk of CVD.^7^ Indeed, those with CKD have a substantially higher chance of dying from CVD than of progressing to end-stage renal disease (ESRD).^8^ Increased arterial stiffness, a marker of CVD risk,^9, 10^ is a commonly recognized feature of CKD,^9^ and an independent predictor of mortality and survival in these patients.^10, 11^ The endothelium is an important regulator of arterial stiffness,^12^ and endothelial dysfunction is also a common feature of CKD^13-15^ and a predictor of CVD.^16^

MicroRNAs (miRs) are small (∼22 nucleotide-long) non-protein coding RNA species involved in post-transcriptional gene product regulation.^17^ In blood, miRs are reported to be stable being protected from degradation by extra-cellular vesicles (such as exosomes), RNA binding protein complexes (such as argonaute 2 – Ago2) and lipoproteins.^18, 19^ As miRs are amplifiable and some are tissue restricted, they represent a new reservoir for biomarker discovery. For example, miR-122 is an established biomarker for liver injury.^20^

miR-126 is enriched in endothelial cells and is a regulator of vascular integrity and angiogenesis. The −3p and −5p forms of miR-126 have activity in endothelial cells – 3p is anti-inflammatory^21^ and 5p is pro-proliferative.^22^ miR-126 is reported to be released from endothelial cells bound to Ago2 and in extra-cellular vesicles, which can transfer functional miR-126 into recipient cells to promote vascular repair.^19^ When endothelial cells were stimulated with the pro-inflammatory cytokine TNFα, the miR-126 cargo of their extra-cellular vesicles was reduced by 80%, consistent with vesicular miR-126 reporting endothelial inflammation.^23^ Reduced levels of circulating miR-126 have been described as a potential biomarker for vascular disorders such as diabetes^24, 25^ and myocardial infarction.^26^ In CKD patients, miR-126 has been reported to fall with worsening renal function.^27^

We hypothesised that circulating miR-126 would report vascular dysfunction in patients with kidney disease. As ‘proof-of-concept’, we first measured miR-126 in a relevant animal model, then in those with AAV. Finally, we measured miR-126 in patients with CKD and ESRD, and explored the relationship with established measures of endothelial function.

## Methods

### Rat model of nephrotoxic nephritis (NTN)

Described elsewhere,^28^ NTN was induced by raising a nephrotoxic serum (NTS) in rabbits to isolated sonicated rat glomeruli which was then injected into male Sprague-Dawley rats (1mL/200 g). A telescoped model of NTN was used where the rats are pre-immunised with 1 mg rabbit immunoglobulin before the injection of NTS.

### Participant groups

#### Patients with AAV

Patients presenting with AAV were recruited at the Royal Infirmary of Edinburgh, UK. Inclusion criteria were seropositivity for ANCA and organ-threatening disease requiring immunosuppression. The study was approved by the local research ethics committee and performed in accordance with the Declaration of Helsinki. Disease activity was graded according to the BVAS (scores range from 0 to 63, with higher scores indicating more active disease)^29^ and by investigators’ assessments of disease activity as remission, ongoing active disease (treatment failure), or relapse. Remission was defined as a BVAS score of 0 that was maintained for 2 months and a prednisone dosage of ≤10 mg/day.

#### Patients with CKD

In brief, subjects were recruited from the renal outpatient clinic at the Royal Infirmary of Edinburgh. The inclusion criteria were: male or female CKD patients, 18-65 years old and clinic BP ≤160/100 mmHg, whether or not on anti-hypertensive medication. We excluded patients with a renal transplant or on dialysis, patients with systemic vasculitis or connective tissue disease, those with a history of established cardiovascular disease, peripheral vascular disease, diabetes mellitus, respiratory disease, neurological disease, current alcohol abuse or pregnancy.

#### Patients with ESRD

Inclusion criteria were: age 18 or over, treated with haemodialysis (HD) for over 3 months. Patients affected by liver disease or with a history of hepato-biliary surgery were excluded; other exclusion criteria were consumption of cytochrome P450-inducing medications, past medical history of epilepsy, cancer, alcoholism and/or psychiatric disease. All patients were treated with HD for 4-5 hours per session, 3 times per week. Data collected included demographic characteristics, cause of ESRD, time on dialysis and current medications. As heparin in blood samples can inhibit polymerase chain reaction (PCR), blood samples were only used when the patient had not been exposed to heparin in the preceding 24 hours.

#### Healthy subjects

Adults with no medical complaints and no medication use were recruited and blood was drawn with informed consent.

### Blood samples

In healthy subjects, blood was collected into 3 EDTA plasma (2.7 ml) and 3 serum (4.9 ml) tubes. One of each type of blood tube was processed without delay – by centrifugation at 1200 × *g* for 10 min at 4^0^C and then supernatant then frozen at −80^0^C. The remaining tubes of blood were left unprocessed at room temperature or 4^0^C for 24h or 7 days. Hemolysis was quantified by spectrophotometric methods as described previously.^30^ In a second study plasma and serum was left at room temperature or 4^0^C for 24h or 7 days.

In patients with AAV, blood samples were taken at study entry (before treatment) and at disease remission (as defined above). For CKD patients, samples were taken from a previously published study and were collected on the study day.^31^ Blood was collected into EDTA tubes and processed immediately as above. Urine was also collected and immediately frozen at −80^0^C.

Plasma asymmetric dimethylarginine (ADMA), an endogenous inhibitor of nitric oxide synthesis, was measured using an optimised, fully validated high performance liquid chromatography method (intra- and inter-assay variations of 1.9% and 2.3%, respectively).^32^ Plasma endothelin-1 (ET-1), the most potent endogenous vasoconstrictor, which contributes to CKD development and progression,^33^ was determined by radioimmunoassay (Peninsular Laboratories Europe, St. Helens, UK) (assay variations 6.3% and 7.2%).^34^ The urate assay was based on the methods of Trivedi and Kabasakalian^35^. Uric acid is oxidized to allantoin by uricase with the production of hydrogen peroxide (H_2_O_2_). The H_2_O_2_ reacts with 4-aminoantipyrine (4-AAP) and 2,4,6-tribromo-3-hydroxybenzoic acid (TBHB) in the presence of peroxidase to yield a quinoneimine dye. The resulting change in absorbance at 548 nm is proportional to the uric acid concentration in the sample. The limits of detection and quantification for the urate assay are 0.01 mmol/l and 0.015 mmol/l, respectively.

### Extra-cellular vesicle isolation

Human plasma was fractionated by differential centrifugation to isolate miR containing proteins and vesicles, as previously described.^36^ Plasma (1 mL) was centrifuged at 500 × *g* for 30 min then 12,000 × *g* for 20 min. The supernatant was then ultracentrifuged at 100,000 × *g* for 1h to pellet extra-cellular vesicles. The remaining supernatant is referred to as the ‘protein fraction’. The vesicles were re-suspended and then pelleted a second time by ultracentrifugation. Extra-cellular vesicle presence and number was quantified by nanoparticle tracking analysis as previously described.^37^ miR concentration in each fraction was determined by PCR, described below.

### Ago2 isolation

MagnaBind goat anti-mouse IgG magnetic bead slurry, 100 μL, (Thermo Scientific, Waltham, USA) was incubated with 10 μg of mouse monoclonal anti-Ago2 (Abcam, Cambridge, UK) or mouse normal IgG (Santa Cruz Biotechnology, Dallas, US) antibodies for 2h at 4°C. The antibody-coated beads were then added to plasma and incubated overnight at 4°C with rotation. Beads were washed and each sample then eluted in RNAse free water before QIAzol was added for RNA isolation. Ago2 isolation was determined by Western blot analysis as described.^38^

### Measurement of arterial stiffness

In a subset of patients gold standard PWV was measured by the foot-to-foot wave velocity method using the SphygmoCor™ system (SphygmoCor™ Mx, AtCor Medical, Sydney, Australia, version 6.31), in which a high-fidelity micromanometer (SPC-301, Millar Instruments, Texas, USA) was used to determine carotid-femoral PWV.^39^

### Measurement of plasma and urine miR

The following miR were measured: miR-126-3p, miR-122-5p, miR-1287 and miR-671.

#### RNA extraction

RNA was extracted from each sample (50 µl) using the miRNeasy serum/plasma kit (Qiagen, Venlo, Netherlands).

#### PCR

After RNA extraction, 5 µl of each eluate was reverse transcribed into cDNA using the miScript II RT Kit (Qiagen, Venlo, Netherlands). The synthesised cDNA was ten-fold diluted and used for cDNA template in combination with the miScript SYBR Green PCR Kit (Qiagen, Venlo, Netherlands) using the specific miScript assays (Qiagen, Venlo, Netherlands). Real-time PCR was performed on a Light Cycler 480 (Roche, Basel, Switzerland) using the recommended miScript cycling parameters. For data analysis (unless stated otherwise) absolute quantification was performed. This used miR mimics for the miRs of interest and running serial dilutions by qRT-PCR. From this, a standard curve was prepared to compare the Ct values for the miRs of interest. This process was carried out for miR-126 and miR-122. In urine Ct values were normalized to miR-39, urinary creatinine or miR-671 (a stable internal normaliser for urine reported by Yang and colleagues^40^).

### Statistical analysis

Data are presented as mean ± standard deviation for patient characteristics and as median and interquartile range (IQR) for all other datasets. Each dataset was analysed for normality using a Shapiro-Wilk test. For non-parametric datasets, comparisons were made using the Mann-Whitney U test or Wilcoxon matched-pairs signed rank test.

## Results

### Circulating miR in NTN model

In NTN rats, plasma miR-126 was reduced compared to untreated controls (**Figure 1**). This reduction in miR-126 was reported by raw Ct values without normalization (control Ct 27.6 (27.0-27.8), n=6, NTN Ct 29.3 (28.4-30.1), n=5, P=0.02) There were no differences in miR-122 or miR-125a-5p between NTN and controls (**Figure 1**).

**Figure 1:**
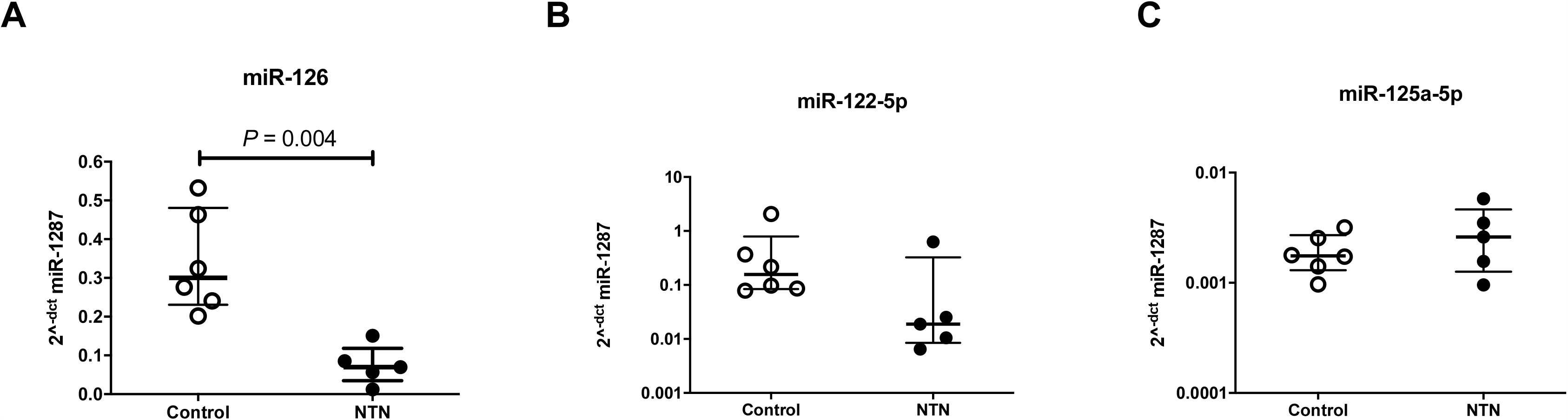
Circulating miRs in rats with nephrotoxic nephritis (NTN) and controls. Each point represents the 2^-dct^ value (normalised by miR-1287) of circulating miR-126 (A), miR-122 (B) and miR-125a (C). The horizontal line represents the median and the bars represent the IQR. Significance determined by Mann-Whitney test.

### Effect of blood processing

In healthy subjects, there was a significant difference in miR-126 concentration between plasma and serum. There was a significant decrease in miR-126 after blood storage for 24h at both room temperature and at 4^0^C (**Supplementary Figure 1a**). After 7d delay in processing, the concentration of miR-126 significantly increased relative to 24h. Hemolysis accompanied this increase in miR-126 (serum - no delay: A_414_ 0.22 (0.20-0.31); 7d delay: A_414_ 1.80 (0.60-1.80), P=0.03. Plasma - no delay: A_414_ 0.16 (0.16-0.28); 7d delay: A_414_ 0.30 (0.22- 0.45), P=0.03). Based on these data only immediately processed plasma samples were used in all subsequent studies. After processing human blood into plasma or serum, storage at room temperature or 4^0^C for 24 hours or 7 days had no statistically significant effect on miR-126 concentration (**Supplementary Figure 1b**). Therefore, after immediate isolation of plasma from blood, samples could be stored for short periods before analysis.

### Circulating miR in AAV

70 patients with AAV were recruited into this study. Pre- and post-treatment clinical data are shown in **Table 1**. Pre-treatment, the plasma miR-126 concentration was 88-fold lower than in healthy controls (AAV median 0.8 fM (IQR 0.3-2.9), healthy subjects 70.4 fM (17.2-770) (**Figure 2**). ROC analysis was performed to quantify the accuracy of miR-126 with regard to distinguishing health from AAV. The area under the curve (AUC) was 0.87 (95%CI 0.80-0.94). At a cut-off of >33fM, specificity was 96% (95%CI 88-99) and sensitivity was 69% (57-80). Post-treatment all 70 patients achieved disease remission (**Table 1**). Plasma miR-126 concentration increased 3.4-fold from pre-treatment levels (post-treatment median 2.7 fM (IQR 0.5-9.8)), but did not return to healthy levels. miR-122 plasma concentrations were not different between pre- and post-treatment samples from patients with AAV (pre-treatment 1.6 fM (0.7-3.4), post-treatment 1.8 fM (0.7-4.7). Urine miR-126 was measured pre- and post- immunosuppressive treatment (**Figure 3**). Using 3 different normalisation methods (spike-in microRNA, urinary creatinine or internal microRNA) the urinary concentration of miR-126 consistently decreased post-treatment.

**Table 1:** Clinical data for AAV patients pre- and post-treatment. The data are shown as mean ± SD with range. Significance of numerical data between groups was ascertained using a 2-tailed paired t-test.

| Characteristics | Pre-treatment (*n* = 70) | Post-treatment (*n* = 70) | P value |
| --- | --- | --- | --- |
| Age, years | 62 ± 14 (27 - 82) | - | - |
| Sex, M/F | 43/27 | - | - |
| Organs involved | 2 ± 1 (1 - 6) | - | - |
| - Kidney | 44 | - | - |
| - Lung | 33 | - | - |
| - ENT | 17 | - | - |
| - Nerve | 13 | - | - |
| - Eyes | 9 | - | - |
| eGFR, mL/min/1.73m<sup>2</sup> | 46 ± 34 (4 - 124) | 57 ± 29 (9 - 121) | 0.04 |
| Creatinine, µmol/L | 200 ± 177 (54 - 962) | 150 ± 144 (64 - 1000) | 0.03 |
| ALT, U/L | 20 ± 15 (6 - 83) | 20 ± 10 (4 - 48) | 0.94 |
| ALP, U/L | 96 ± 62 (13 - 338) | 67 ± 18 (39 - 122) | <0.001 |
| GGT, U/L | 55 ± 44 (11 - 182) | 33 ± 26 (10 - 148) | <0.001 |
| Albumin, g/L | 28 ± 7 (14 - 41) | 37 ± 4 (24 - 46) | <0.0001 |
| CRP, mg/L | 82 ± 75 (3 - 275) | 6 ± 15 (0 - 108) | <0.0001 |
| Hemoglobin, g/L | 107 ± 23 (62 - 157) | 124 ± 28 (2 - 170) | <0.0001 |
| Urine protein:creatinine mg/mmol | 141 ± 188 (0 - 948) | 107 ± 146 (0 - 531) | 0.82 |

**Figure 2:**
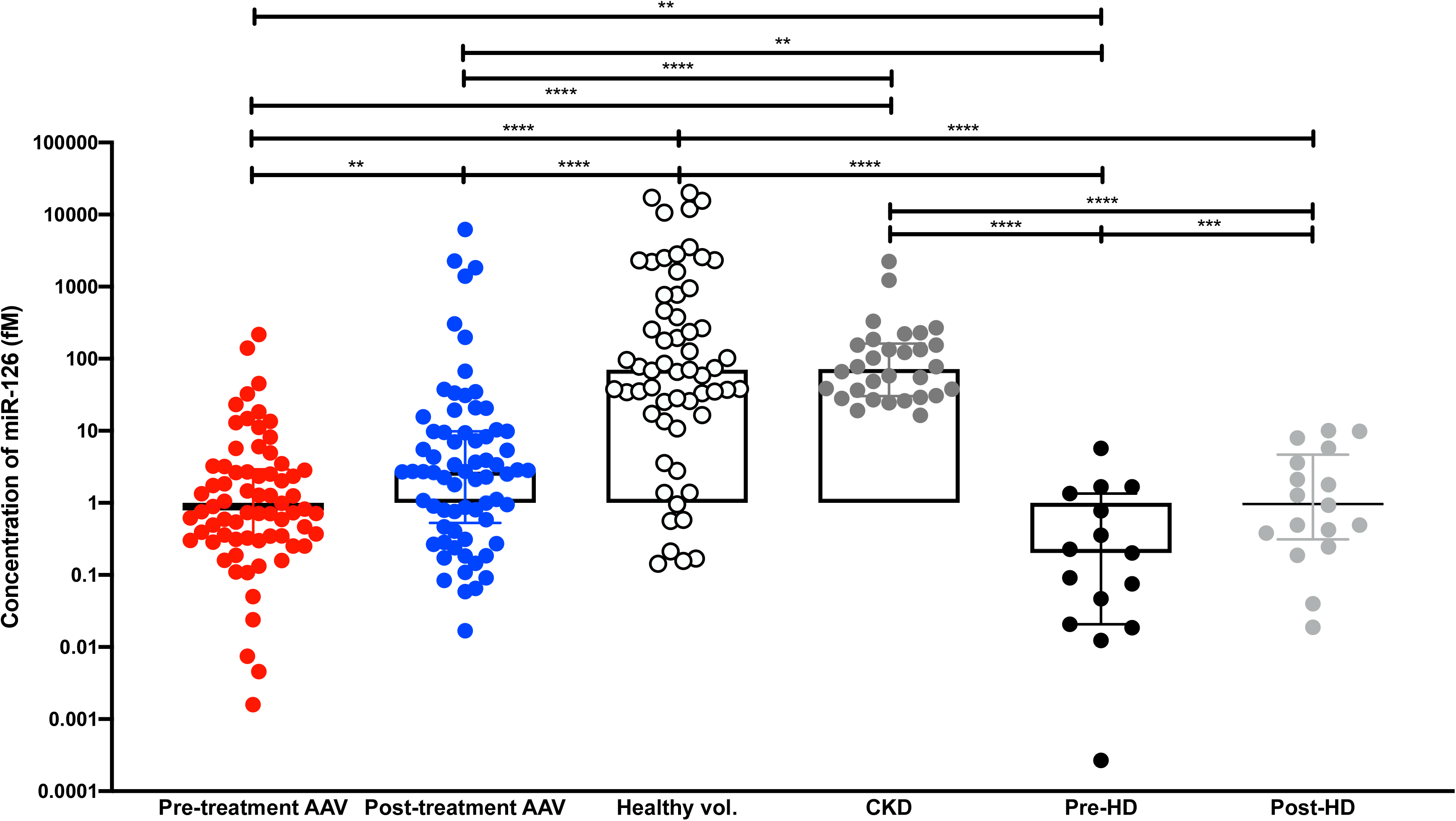
The concentration of miR-126 in ANCA-associated vasculitis (AAV) patients (n = 70 paired) pre- and post-treatment, healthy volunteers (n = 60), chronic kidney disease (CKD) (n=30) and end-stage renal disease (n=15 paired) pre- and post-hemodialysis (HD). Bars show median with IQR, **p< 0.01, ****p<0.0001 (Mann-Whitney test for unpaired analysis and Wilcoxon test for paired analysis).

**Figure 3:**
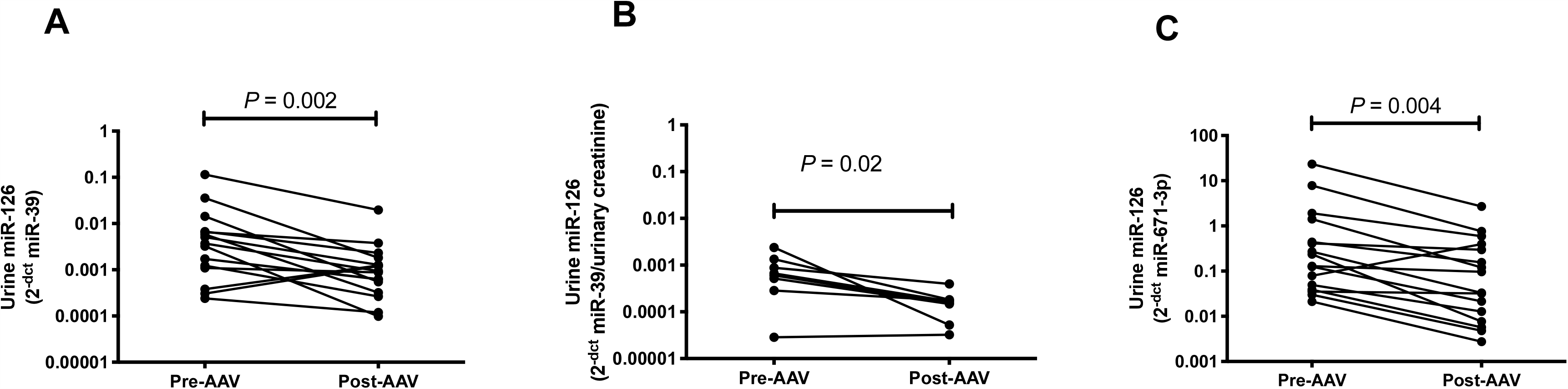
Urine miR-126 in patients with AAV, pre- (Pre-AAV) and post- (Post-AAV) treatment. Each point represents the 2^-dct^ value (normalised by miR-39 spike-in control (A), urinary creatinine concentration (B) and miR-671 (C)) of miR-126.

### Mechanism of miR-126 release into the circulation

miRs circulate bound to proteins or encapsulated in extra-cellular vesicles such as exosomes. Plasma from healthy volunteers was fractionated by differential centrifugation and extra-cellular vesicles were isolated (confirmed by nanoparticle tracking analysis – **Figure 4A**). miR-126 was enriched in the ‘protein fraction’ as opposed to the extra-cellular vesicle fraction (29-fold increase in supernatant compared to exosome pellet) (**Figure 4B**). The RNA binding protein Ago2 was isolated from plasma and the bound miR-126 concentration measured. There was a significant increase in Ago2-bound miR-126 in those with AAV post-treatment (**Figure 4C**), consistent with this miR biomarker being specifically bound to Ago2. By contrast there was no significant treatment-induced change in miR-126 in the extra-cellular vesicle fraction (**Figure 4D**). There was no increase in Ago2-bound miR-122 and no change in plasma Ago2 concentration with treatment (data not shown).

**Figure 4:**
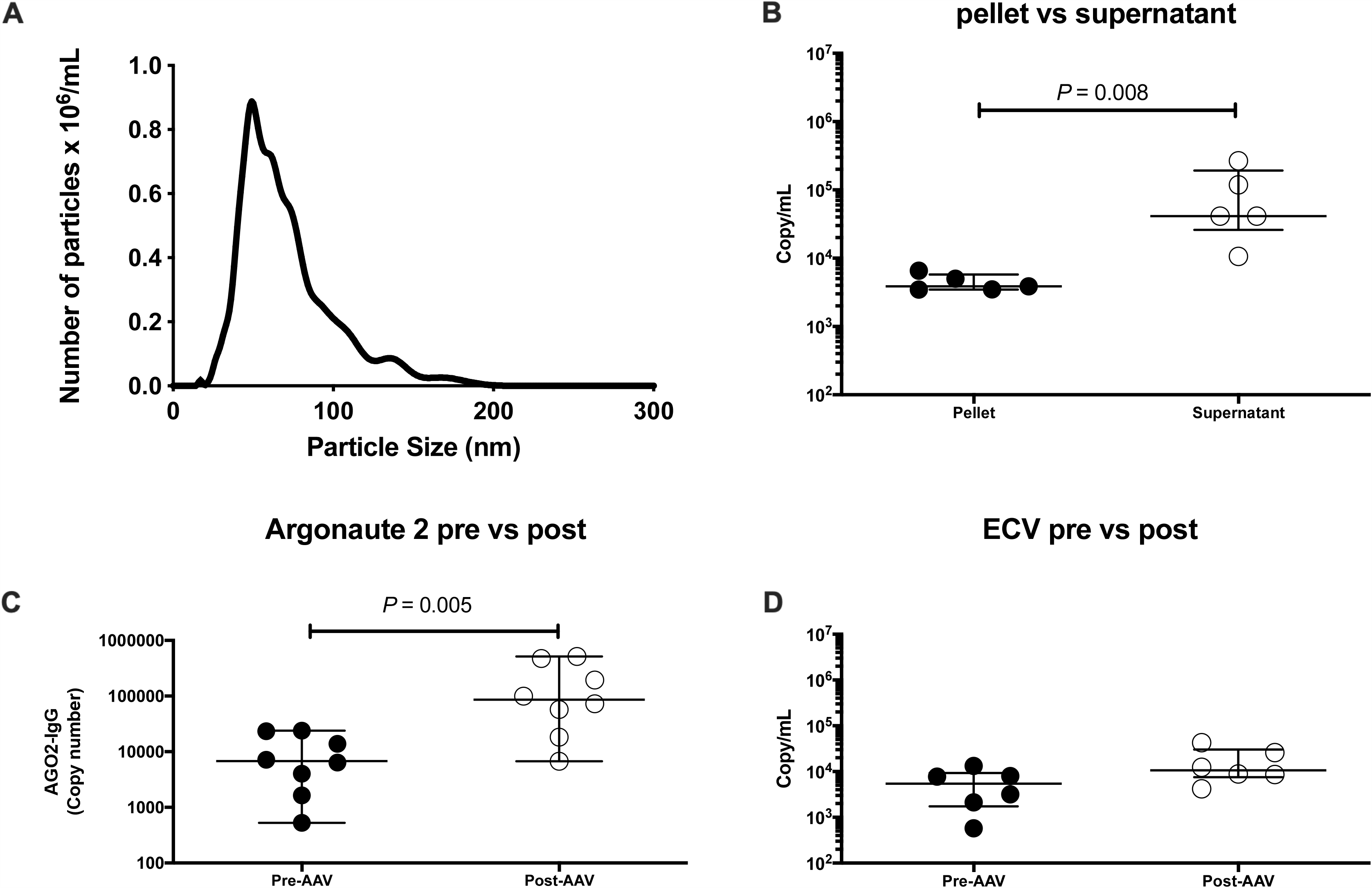
miR-126 is bound to argonaute 2 (Ago2) in human plasma. (A) size and number of particles measured by nanoparticle tracking analysis (NTA) following isolation of extra-cellular vesicles from human plasma by differential centrifugation. (B) Dot plot of miR-126 measured in the extra-cellular vesicle containing pellet and supernatant after ultracentrifugation of human plasma. (C) Dot plot of miR-126 measured in the antibody isolated Ago2 fraction from 8 AAV patients before treatment (Pre-AAV) and after treatment (Post-AAV). The y-axis represents copy number obtained from the Ago2 pull-down minus the copy number obtained from IgG control pull-down from the same sample. (D) Dot plot of miR-126 measured in the extra-cellular vesicle containing pellet from 8 AAV patients before treatment (Pre-AAV) and after treatment (Post-AAV). The horizontal line represents the median and the error bars represent the IQR.

### Circulating miR-126 in CKD

Having demonstrated proof-of-concept that miR-126 changes with acute endothelial/kidney injury, we explored its circulating concentration in CKD. This condition is associated with chronic endothelial dysfunction and high cardiovascular risk. Samples were analysed from patients with CKD (n=30) and hemodialysis-dependent ESRD (n=15), immediately before and after dialysis (**Table 2**). In patients with ESRD, miR-126 was substantially reduced compared with healthy subjects and patients with CKD (352- and 358-fold, respectively). There was a modest increase following a hemodialysis session (4.7-fold) (**Figure 2**). There was a range of plasma miR concentrations in patients with CKD, which we hypothezised might reflect differences in vascular health across this patient group. For these patients we correlated miR concentrations against a range of well-recognized measures of vascular function. miR-126 correlated significantly with PWV, circulating ADMA, ET-1 and urate concentrations as well as proteinuria (**Figure 5**). miR-126 correlated weakly with serum creatinine (r^2^=0.12). There was no relationship between miR-122 and any of the measures of vascular or renal function assessed.

**Table 2:** Clinical data obtained for CKD patients and ESRD patients pre-hemodialysis (HD). The data are shown as mean ± SD with range.

| **Characteristics** | **CKD (*n* = 30)** | **Pre-HD (*n* = 15)** |
| --- | --- | --- |
| **Age, years** | 56 ± 16 (26 - 82) | 59 ± 12 (34 - 82) |
| **Sex, M/F** | 21/9 | 8/7 |
| **eGFR, mL/min/1.73m<sup>2</sup>** | 25 ± 14 (7 - 57) | - |
| **Creatinine, µmol/L** | 217 ± 133 (76 - 654) | - |
| **ALT, U/L** | 19 ± 8 (9 to 50) | 15 ± 6 (7 - 25) |
| **ALP, U/L** | 97 ± 38 (40 - 182) | 100 ± 43 (52 to 178) |
| **CRP, mg/L** | 5 ± 2 (2 - 7) | 14 ± 21 (2 to 77) |
| **Haemoglobin, g/L** | 123 ± 16 (83 - 160) | 122 ± 12.51 (94 - 139) |

**Figure 5:**
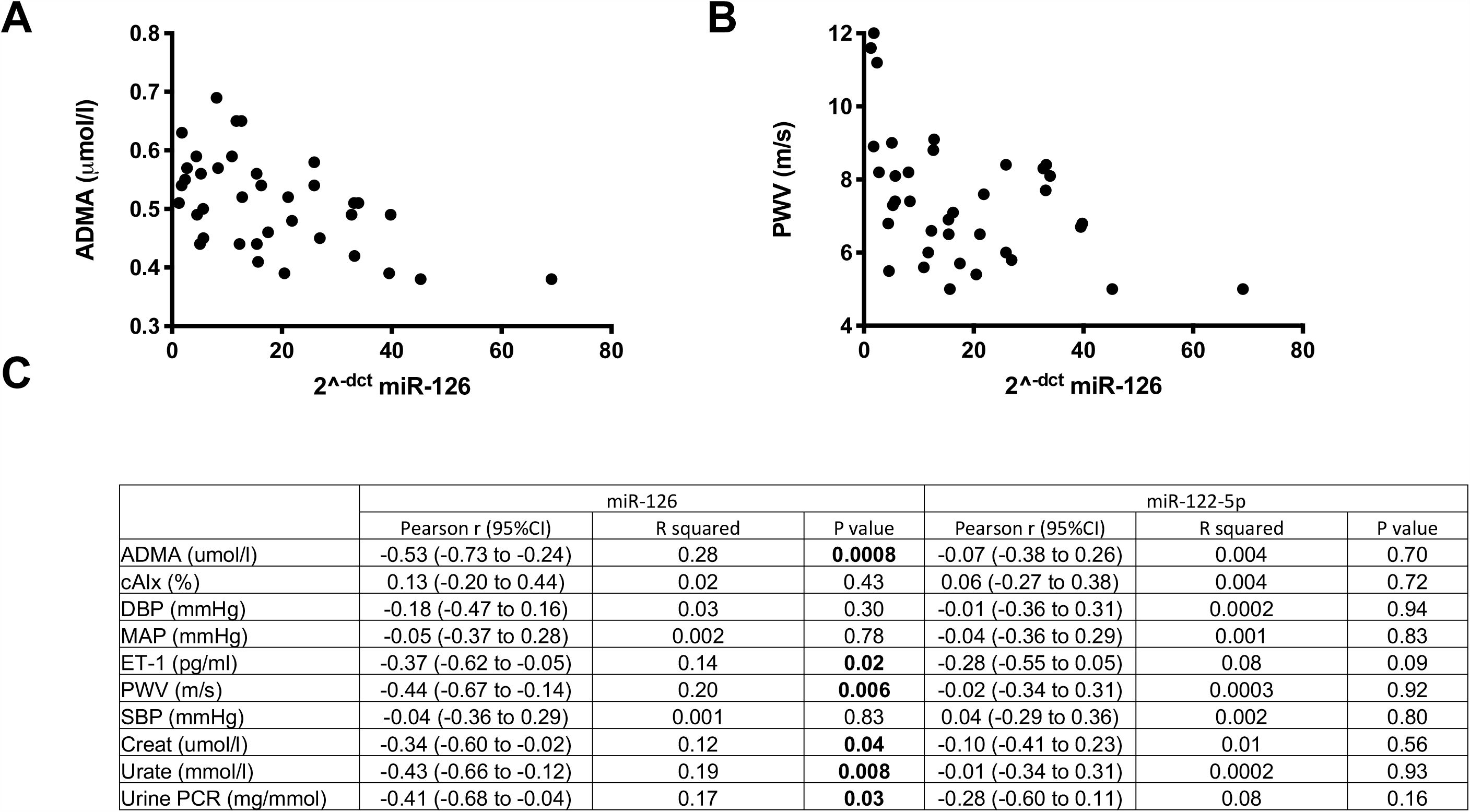
Scatter plots of asymmetric dimethylarginine (ADMA) (A) and pulse wave velocity (PWV) (B) versus circulating miR-126 in CKD patients. (C) Table with Pearson r (95% CI), r^2^ and *P* values of correlation between miR-126 and miR-122-5p and other biomarkers.

## Discussion

The current study develops the potential role of miR-126 as a circulating biomarker of vascular function in acute and chronic kidney disease. We initially achieved proof of concept in rodents and then went on to study AAV as a prototype of severe, acute kidney injury and vascular dysfunction that is aggressively treated with immunosuppression. Then, we went on to study patients with CKD as they have a heavy burden of vascular dysfunction. In a clinically-relevant rat model, a lower circulating miR-126 (but not other miR species) was associated with disease. We have demonstrated that in patients with AAV, circulating miR-126 concentrations are low at disease presentation and rise with successful treatment. Pre-treatment levels are lower than those seen in health and in CKD. Following treatment, miR-126 rises but not to levels seen in health. A liver specific miR species (miR-122) showed no differences in AAV patients before or after treatment. In patients with CKD, miR-126 correlated with other markers of vascular health. miR-126 was substantially reduced in patients with ESRD. Thus, miR-126 may be a useful marker of vascular inflammation and, with development, may help clinical decision-making.

miRs are an area of substantial research interest, in part because they represent potential disease biomarkers. It is widely believed that one of their key properties is that they are stable in the circulation.^18^ In the current study we have demonstrated that for miR-126 this is not the case: significant degradation occurred if the sample was left unprocessed for 24h. We have reported similar instability for miR-122.^41^ We also demonstrated that hemolysis may increase the concentration of miR-126 present in the sample. Furthermore, despite reports that miRs are detected at similar concentrations in plasma and serum, plasma miR-126 was substantially higher than serum. Therefore, we recommend the immediate processing of plasma samples in future studies of miR-126.

miR-126 increased in the circulation following successful treatment of patients with AAV. Given that there was no change in miR-122, the increase in circulating miR-126 is unlikely to reflect a global reduction in miR excretion or a non-specific increase in miR expression. In spot urine samples, miR-126 was reduced by treatment, which may contribute to the increase in the circulation. miR circulate encapsulated in extra-cellular vesicles such as exosomes and bound to proteins especially Ago2, but the relative contribution and biological importance of each fraction is controversial.^36^ In our study, when plasma was fractionated by centrifugation miR-126 predominately remained in the supernatant. This is consistent with studies that report a low amount of miR-126 in human exosomes.^36^ Importantly, we demonstrate here that miR-126 is bound to the circulating protein Ago2 and this fraction increases in patients with AAV following successful disease treatment. It is reported that stimulation of endothelial cells with TNFα resulted in a decrease of miR-126.^23^ This is in keeping with our own findings and suggests that the reduction in circulating miR-126 in those with active AAV may reflect a degree of endothelial dysfunction secondary to inflammation. This hypothesis is further supported by miR-126 being enriched in the endothelium^21^ and our strong correlation between the circulating miR-126 and markers of endothelial function such as pulse wave velocity and ADMA. From a clinical perspective, a circulating biomarker that reports vascular health could have widespread utility and prospective studies should qualify miR-126 in a range of settings, including other systemic inflammatory disorders such as rheumatoid arthritis and systemic lupus erythematosus.

In keeping with our hypothesis that miR-126 might act as a measure of vascular integrity, circulating levels did correlate with well-recognized measures of endothelial dysfunction in our CKD cohort namely, high PWV, ADMA, ET-1, and urate. Plasma ADMA, an endogenous inhibitor of nitric oxide synthase, and plasma ET-1 were measured as components of the nitric oxide and ET systems, respectively. Both contribute to vascular dysfunction in CKD and an imbalance (more ET-1/ less nitric oxide) may contribute to vasoconstriction, inflammation and atherosclerosis.^33, 42^ Serum urate has also emerged as an important risk factor for cardiovascular risk and CKD progression.^43^ Treatment of asymptomatic hyperuricemia has been shown to improve renal function^44^ and delay disease progression^45^ in patients with early CKD. From a clinical perspective, ET receptor antagonism is being investigated as a novel therapeutic strategy for renoprotection in CKD.^46^ It has been shown to not only lower proteinuria but also serum urate.^47, 48^ Furthermore, given there is often reciprocal up-regulation of the nitric oxide system when the ET system is down-regulated^33^ an ET blocking strategy may offset some of the potentially deleterious effects of elevated circulating ADMA.

Interestingly, our data suggest no substantial difference in circulating miR-126 between health and CKD. The lack of a clear difference may relate to the CKD population we studied as they had minimal comorbidity without overt CVD. Additionally, we excluded those with diabetes. Indeed, the ‘vascular health’ of our CKD subjects is demonstrated by a mean pulse wave velocity of 6.8 m/s, significantly lower than that of 8.2 m/s for a group with a similar eGFR reported in a study by Wang *et al*.^49^ They used a similar technique for measuring arterial stiffness but included patients with diabetes and CVD. Furthermore, our subjects were relatively young (∼50 years), with good blood pressure control (∼135/80 mmHg) and reasonably preserved renal function (eGFR ∼75 ml/min). Thus, future studies should extend to the wider spectrum of CKD.

With further clinical development miR-126 may have utility as a biomarker of vascular dysfunction in patients with kidney disease (and possibly in people with normal kidney function). Such a miR biomarker may provide a read out of cardiovascular drug efficacy that translates from pre-clinical models into early phase clinical trials. In support of this, we have demonstrated that miR-126 tracks effective treatment in patients with AAV and reports injury when translated back into a rat model of nephrotoxic nephritis. The work presented here provides proof of concept data to build on in larger studies that test utility, both in the specific clinical scenario of reporting disease activity in vasculitis, and across a broader group of patients with acute kidney injury and CKD.

## Data Availability

Data available on request.

## Acknowledgments

Author JWD acknowledges the support of NHS Research Scotland (NRS) through NHS Lothian and a BHF Centre of Research Excellence Award.

## Source of Funding

Author KS was funded by a British Heart Foundation PhD Studentship FS/15/63/32033. Author ADBV was supported by an NC3Rs PhD Studentship (NC/K001485/1). TF is supported by a Clinical Research Training Fellowship from the *Medical Research Council* (MR/R017840/1). Author ND was supported by a British Heart Foundation Intermediate Clinical Research Fellowship (FS/13/30/29994). RWH is supported by a Wellcome Clinical Research Career Development Fellowship (209562/Z/17/Z).

## Disclosures

None of the authors have any conflicts of interest.

## Legends

**Supplementary figure 1:** A) Bar graphs displaying median Ct values of miR-126 measured by qPCR. Blood samples were collected from healthy volunteers (n=6). One serum and plasma blood tube was centrifuged and the supernatant frozen at −80 ^0^C without delay. The remaining blood tubes were left at room temperature (RT) or 4^0^C for 24h or 7 days before being processed and stored at −80^0^C. B) Serum and plasma samples were immediately centrifuged to isolate serum/plasma. One serum and one plasma sample were stored at −80^0^C without delay and the remaining tubes were left at room temperature (RT) or 4^0^C for 24h or 7 days. Error bars represent the interquartile range.

## Notes

### Competing Interest Statement

The authors have declared no competing interest.

### Author Declarations

South-East Scotland Research Ethics Committee

